# Increased insulin resistance due to Long COVID is associated with depressive symptoms and partly predicted by the inflammatory response during acute infection

**DOI:** 10.1101/2022.12.01.22283011

**Authors:** Hussein Kadhem Al-Hakeim, Haneen Tahseen Al-Rubaye, Abdulsahib S. Jubran, Abbas F. Almulla, Shatha Rouf Moustafa, Michael Maes

**Affiliations:** Department of Chemistry, College of Science, University of Kufa, Iraq; College of Medical Laboratory Techniques, Imam Ja’ afar Al-Sadiq University, Najaf, Iraq; University of Alkafeel, College of Dentistry, Najaf, Iraq; Medical Laboratory Technology Department, College of Medical Technology, The Islamic University, Najaf, Iraq; Clinical Analysis Department, College of Pharmacy, Hawler Medical University, Havalan City, Erbil, Iraq; Department of Psychiatry, Faculty of Medicine, Chulalongkorn University, Bangkok, Thailand; Department of Psychiatry, Medical University of Plovdiv, Plovdiv, Bulgaria; Deakin University, IMPACT, the Institute for Mental and Physical Health and Clinical Translation, School of Medicine, Barwon Health, Geelong, Australia

**Author notes:** Corresponding Author: Prof. Dr. Michael Maes, M.D., Ph.D., Department of Psychiatry, Faculty of Medicine, King Chulalongkorn Memorial Hospital, Bangkok, Thailand,. (6) Michael Maes (researchgate.net), Michael Maes - Google Scholar.

**Keywords:** inflammation, Long-COVID, oxidative and nitrosative stress, major depression, mood disorders, biomarkers

## Abstract

**Background:** Some months after the remission of acute COVID-19 infection, some people show depressive symptoms, which are predicted by increased peak body temperature (PBT) and lowered blood oxygen saturation (SpO2). Nevertheless, no data indicate whether Long COVID is associated with increased insulin resistance (IR) in association with depressive symptoms and immune, oxidative, and nitrosative (IO&NS) processes.

**Methods:** We used the homeostasis Model Assessment 2 (HOMA2) calculator^©^ to compute β-cell function, insulin sensitivity and resistance (HOMA2-IR) and measured the Beck Depression Inventory (BDI) and the Hamilton Depression Rating Scale (HAMD) in 86 Long COVID patients and 39 controls. We examined the associations between the HOMA2 indices and PBT and SpO2 during acute infection, and depression, IO&NS biomarkers (C-reactive protein, NLRP3 activation, myeloperoxidase, and advanced oxidation protein products) 3-4 months after the acute infection.

**Results:** Long COVID is accompanied by increased HOMA2-IR, fasting blood glucose, and insulin levels. We found that 33.7% of the patients versus 0% of the controls had HOMA2-IR values >1.8, suggesting IR. PBT, but not SpO2, during acute infection significantly predicted IR, albeit with a small effect size. Increased IR was significantly associated with depressive symptoms as assessed with the BDI and HAMD above and beyond the effects of IO&NS pathways. There were no significant associations between increased IR and the activated IO&NS pathways during Long COVID.

**Conclusion:** Long COVID is associated with new-onset IR in a subset of patients. Increased IR may contribute to the onset of depressive symptoms due to Long COVID by enhancing overall neurotoxicity.

## Introduction

The pathogenesis of acute COVID-19 includes SARS-CoV-2 entry into the host respiratory epithelial cells followed by viral translation and multiplication in the cytoplasm and infection of nearby host cells ^(1-4)^. These processes are accompanied by the activation of immune-inflammatory pathways and may result in pneumonia, lung injury, and excessive inflammatory reactions including a cytokine storm that may cause disseminated intravascular coagulation and multisystem failure ^(1-4)^. SARS-CoV-2 may activate the nucleotide-binding domain, leucine-rich repeat, and pyrin domain-containing protein 3 (NLRP3) inflammasome with elevations in interleukin (IL)-1β, IL-18, and caspase 1^(1-5)^.

After remission of the acute phase of COVID-19, many people experience Long COVID which comprises a variety of neuropsychiatric symptoms, such as neurocognitive impairments, sleep disturbances, affective symptoms (low mood and anxiety), chronic fatigue, and somatic symptoms like dyspnea, autonomic symptoms, and pain symptoms ^(6-15)^. Within six months of the onset of COVID-19 symptoms, over one-third of COVID-19 survivors may suffer from such neuropsychiatric symptoms ^(16)^.

Recently, we found that both acute and long-term COVID are characterized by increases in a) fatigue and physiosomatic symptoms such as headache, malaise, fibromyalgia-like, gastrointestinal, cardiovascular, and autonomous symptoms, b) anxiety symptoms including anxious mood, tension, irritability, and fears, and c) depressive symptoms such as low mood, feelings of guilt, and loss of interest ^(17-20)^. Moreover, in both acute and Long-COVID, one factor (latent vector) could be extracted from these diverse neuropsychiatric symptoms, suggesting that acute and Long-COVID are characterized by a wide range of neuropsychiatric symptoms which are driven by a single latent characteristic, named the “physio-affective phenome” ^(17-20)^.

Important predictors of the severity of the physio-affective phenome of acute and Long COVID are lowered oxygen saturation (SpO2) and an increased peak body temperature (PBT) during the acute phase ^(20)^. Both lowered SpO2 and increased PBT are sensitive markers of the intensity of the immune-inflammatory response during acute COVID-19 and predict not only critical disease and death due to COVID-19, but also the physio-affective phenome of both acute and Long COVID ^(20-22)^.

Moreover, activated immune-inflammatory, oxidative, and nitrosative stress (IO&NS) pathways are significantly associated with the physio-affective symptoms of Long COVID ^(17, 18)^. In different studies, we observed that activation of the NLRP3 inflammasome (as indicated by alterations in interleukin (IL)-1β, IL18, and caspase 1), increased C-reactive protein (CRP), myeloperoxidase (MPO), malondialdehyde (MDA), protein carbonyls, and advanced protein oxidation products (AOPP) and lowered zinc and glutathione peroxidase (Gpx) are associated with Long COVID’ physio-affective symptoms ^(18)^.

People with COVID-19 exhibit an increased risk and excess burden of incident diabetes ^(23)^. For example, glycaemic abnormalities could be detected two months after recovery from COVID-19 ^(24)^. Mood disorders including major depression and anxiety disorders are not only accompanied by activated IO&NS pathways ^(25, 26)^ but also by relative insulin resistance (IR) and a significant association between IR assessments and depressive symptoms ^(26-30)^. The neuro-immune-toxicity theory of mood disorders conceptualizes that increased neurotoxicity due to multiple neurotoxic IO&NS pathways and neurotoxicity due to IR are involved in the pathophysiology of mood disorders ^(25-27)^. Importantly, in mood and anxiety disorders, increased damage due to O&NS, as indicated by increased AOPP and MDA levels, is significantly associated with increased IR, suggesting that, in mood disorders, increased O&NS plays a role in the onset of IR ^(26, 27)^. Furthermore, increased lipid peroxidation, aldehyde production, and chlorinative stress are characteristics of the metabolic syndrome (MetS) and are known to mediate IR and atherogenicity in MetS. Inflammatory pathways and the NLRP3 inflammasome play an important role in the pathogenesis of IR, type 2 diabetes mellitus (T2DM), and obesity ^(31, 32)^. Nevertheless, there are no studies that have examined whether the physio-affective symptoms of Long COVID are associated with increased IR and whether the latter is associated with signs of activated IO&NS pathways during the acute infectious and Long COVID phases.

Hence, the present study aimed to examine a) whether Long COVID is associated with increased IR; b) whether increased IR in Long COVID is associated with the physio-affective phenome of Long COVID, the inflammatory response of the acute infectious phase (as indicated by increased PBT and lowered SpO2) and activated IO&NS pathways (as indicated by increased CRP, NLRP3, AOPP, and MPO levels); and d) IR impacts the physio-affective phenome above and beyond the effects of these IO&NS biomarkers.

## Participants and Methods

### Participants

In the present study, we combined a case-control research methodology with a retrospective cohort study design to assess the impact of acute phase biomarkers on the IR parameters in Long COVID patients. The case-control strategy allowed us to analyze differences between controls and Long COVID subgroups. In the last three months of 2021, we included 86 individuals who had at least two Long COVID symptoms and had previously been identified and treated for an acute COVID-19 infection. The patients were identified using the officially published WHO criteria for post-COVID (long COVID)^(33)^, which include the following: a) a history of proven SARS-CoV-2 infection; b) symptoms that persisted beyond the acute stage of illness or that manifested during recovery from acute COVID-19 infection, lasted at least two months, and were present 3–4 months after the onset of COVID-19; and c) patients who have at least 2 symptoms ^(33)^.

All patients were admitted to one of the official quarantine facilities in the city of Al-Najaf that specializes in the treatment of acute COVID-19, including the Hassan Halos Al-Hatmy Hospital for Transmitted Diseases, the Middle Euphrates Center for Cancer, the Imam Sajjad Hospital, the Al-Hakeem General Hospital, and the Al-Zahraa Teaching Hospital for Maternity and Pediatrics. During the acute infectious phase, all patients included here showed: a) acute respiratory syndrome and the disease’s typical symptoms of fever, breathing problems, coughing, and loss of smell and taste; b) positive reverse transcription real-time polymerase chain reaction (rRT-PCR) findings; and c) the presence of positive SARS-specific IgM antibodies. During Long COVID these patients had negative rRT-PCR results after the acute period. We also included 39 controls, who were either staff members or members of their families or friends, and selected from the same catchment area. All 39 controls had negative rRT-PCR results and showed no clinical signs of an acute infection, such as a dry cough, sore throat, shortness of breath, fever, night sweats, and chills. Exclusion criteria for controls were a lifetime and current history of psychiatric axis-1 disorders, such as major affective disorders (major depressive disorder and bipolar disorder), dysthymia, generalized anxiety disorder, panic disorder, schizo-affective disorder, schizophrenia, psycho-organic syndrome, substance use disorders—with the exception of tobacco use disorder—and chronic fatigue syndrome and fibromyalgia. Nevertheless, to account for the confounding effects of psychological stress due to the COVID-19 pandemic (including the effects of lockdown and social isolation), we allowed that around one-third of the controls would show some distress or adjustment symptoms. Consequently, some controls (one-third) showed Hamilton Depression Rating Scale (HAMD) scores ^(34)^ between 7 and 12. Long COVID patients were excluded for a lifetime history of the same disorders as described for controls and a current diagnosis of bipolar disorder, schizo-affective disorder, schizophrenia, psycho-organic syndrome, and substance use disorders (with the exception of tobacco use disorder). Additionally, we excluded patients and controls who had T2DM and T1DM, systemic (auto)immune diseases like inflammatory bowel disease, psoriasis, scleroderma, rheumatoid arthritis, liver or renal disease, as well as neurodegenerative and neuroinflammatory disorders including Parkinson’s and Alzheimer’s disease, multiple sclerosis, or stroke. Moreover, according to the requirements of the HOMA calculator software, the following patient exclusion criteria applied: patients with obvious serious overt diabetes complications, such as heart illnesses, liver diseases, and renal diseases; and patients whose fasting insulin was >400 pM. Additionally, since metformin may have an impact on insulin sensitivity and IR ^(35)^, we did not include individuals who were using metformin ^(36)^. Women who were pregnant or nursing were also excluded from this research.

Before participating in the study, all controls and patients, or their parents or legal guardians, supplied written informed consent. The Najaf Health Directorate, Training and the Human Development Center (Document No. 18378/2021) and the institutional ethics boards of the University of Kufa (8241/2021) both gave their approval for the study. The International Guideline for Human Research Safety is followed by our institutional review board, and the study was carried out in accordance with Iraqi and international ethical and privacy laws, such as the World Medical Association’s Declaration of Helsinki, The Belmont Report, the CIOMS Guideline, and the International Conference on Harmonization of Good Clinical Practice (ICH-GCP).

#### Measurements

Three to four months after the acute infectious phase of COVID-19 (mean ± SD duration of illness: 14.68 ± 5.33 weeks), a senior psychiatrist used a semi-structured interview to collect sociodemographic and clinical data from controls and Long COVID patients. Depression severity was evaluated using the HAMD ^(34)^ and the Beck-Depression Inventory (BDI-II) ^(37)^ scale scores. The diagnosis of TUD was made using DSM-5 criteria. We recorded the immunizations received by the participants, namely AstraZeneca, Pfizer, or Sinopharm. The body mass index (BMI) was calculated by dividing the body weight in kilograms by the square of the individual’s height in meters.

A well-trained paramedical professional measured SpO2 using an electronic oximeter (Shenzhen Jumper Medical Equipment Co., Ltd.), and body temperature with a digital thermometer (sublingual until the beep). The lowest SpO2 and PBT readings recorded during the acute period of illness were extracted from patient records. We created a new indicator based on these two evaluations that represent decreased SpO2 and increased PBD as the z transformation of the latter (z PBT) - z SpO2 (dubbed zPBT-zSPO2).

Fasting blood samples were collected in the early morning between 7.30 and 9 a.m., before having breakfast. Five milliliters of venous blood were extracted and transferred to clean simple tubes. Hemolyzed specimens were discarded. After fifteen minutes, the blood samples were centrifuged at 3000 rpm for five minutes to separate the serum, which was then transferred to three fresh Eppendorf tubes for testing. Fasting Blood Glucose (FBG) levels were tested spectrophotometrically using a kit provided by Biolabo^®^ (Maizy, France). Commercial ELISA sandwich kit were used to measure serum insulin (DRG^®^ International Inc., USA). The sensitivity of the insulin assay was 1.76 μIU/ml (12.22pM) and the CV% was 2.6%. Homeostasis Model Assessment 2 (HOMA2) calculator^©^ (Diabetes Trials Unit, University of Oxford; https://www.dtu.ox.ac.uk/homacalculator/download.php) was used to calculate β-cell function (HOMA2%B), insulin sensitivity (HOMA2%S), and IR (HOMA2-IR) from the fasting serum insulin and glucose levels. We used 2 threshold values of HOMA2-IR to denote IR (HOMA2-IR>1.8) and MetS (HOMA2-IR>1.4) ^(38)^. Furthermore, we computed two new z unit-based composite scores, namely, a first reflecting IR as zFBG+zInsuline (zFDG+zINS), and a second reflecting β-cell function computed as zINS-zFBG ^(26)^. In the current study, zFBG+zINS was significantly correlated with the HOMA2-IR index (r=0.892, p<0.001, n=125) and zINS-zFBG with HOMA2%B (r=0.987, p<0.001, n=125). We employed the C-Reactive Protein (CRP) latex slide test (Spinreact®, Barcelona, Spain) to assay CRP serum levels. ELISA kits from Nanjing Pars Biochem Co., Ltd. (Nanjing, China) were used to quantify serum levels of MPO, AOPP, IL-1β, IL-18, and caspase-1. The intra-assay CV values for all assays were less than 10%. Consequently, we computed different composite scores, namely reflecting a) oxidative stress toxicity (OSTOX) computed as the sum of z AOPP + z MPO; b) NLRP3 computed as z IL-1β + z IL-18 + z caspase 1; c) inflammation as z IL-1β + z IL-18 + z caspase + z CRP (labeled: zNLPR3+zCRP) ^(18)^; and d) an overall neurotoxicity index as zNLRP3+zCRP+zOSTOX+zIR.

#### Statistical analysis

Analysis of variance (ANOVA) or the Kruskal-Wallis test (in case of heterogeneity of variance) was used to examine differences in scale variables across groups, whilst analysis of contingency tables (χ^2^-test) was employed to examine the relationships between nominal variables. To evaluate relationships between biomarkers and clinical ratings, we generated correlation matrices using Pearson’s product-moment correlation coefficients. Using the univariate generalized linear model (GLM) approach, we determined the relationships between diagnosis and biomarkers while adjusting for confounding factors such as TUD, age, sex, and BMI. We derived estimated marginal mean (SE) values produced by the GLM. Using multiple regression analysis, the significant biomarkers that predict physio-affective assessments were identified. We also used an automated stepwise procedure with a p-value of 0.05 to enter and p=0.06 to remove. We determined the standardized beta coefficients for each significant explanatory variable using t statistics with accurate p-values, model F statistics, and total variance explained (R^2^). In addition, we investigated the residual plots and checked for multicollinearity utilizing the variance inflation factor (VIF) and tolerance, and using the White and modified Breusch-Pagan tests, we tested for heteroskedasticity. K means cluster analysis was performed to delineate a subgroup of patients with aberrations in the biomarkers and increased depression rating scales as well. All statistical analyses were conducted using version 28 of IBM SPSS for Windows.

Partial least squares (PLS) analysis ^(39)^ was used to determine a) the associations between biomarkers entered as input variables and symptom domains scores entered as output variables. Data were entered as either single indicators (all biomarkers) or a latent vector generated from the symptom dimensions. PLS path analysis was conducted using 5,000 bootstrap samples only when: a) the overall quality of the model as indicated by Standardized Root Mean Squared Error (SRMR) 0.080 was adequate; b) the latent vectors extracted from indicators had adequate reliability as indicated by average variance extracted (AVE) > 0.500, Cronbach alpha > 0.7, composite reliability > 0.7, rho_A > 0.80; c) all indicators loaded highly (> 0.660) at p<0.001 on the latent vector; and d) construct cross-validated redundancies are adequate ^(39)^. We employed complete bootstrapping (5,000 subsamples) and PLS path modelling to compute path coefficients with p-values and total and indirect effects ^(40)^.

## Results

### Sociodemographic data and IR measurements

**Table 1** shows the sociodemographic data and IR measurements in healthy controls and patients with Long COVID. There were no significant differences in age, sex, BMI, married status, urban/rural distribution, vaccination status, and smoking between both groups, although education was marginally higher in the patient group. The total BDI and HAMD scores were significantly higher in patients than in controls. FBG, insulin, HOMA2-IR, zFBG+zINS were significantly higher in patients than in controls, whereas HOMA2%S and zINS-zFBG were significantly lower in patients. The incidence of IR (defined as HOMA2-IR > 1.8) and MetS (defined as HOMA2-IR >1.4) were significantly higher in Long COVID patients than in healthy controls.

**Table 1.**
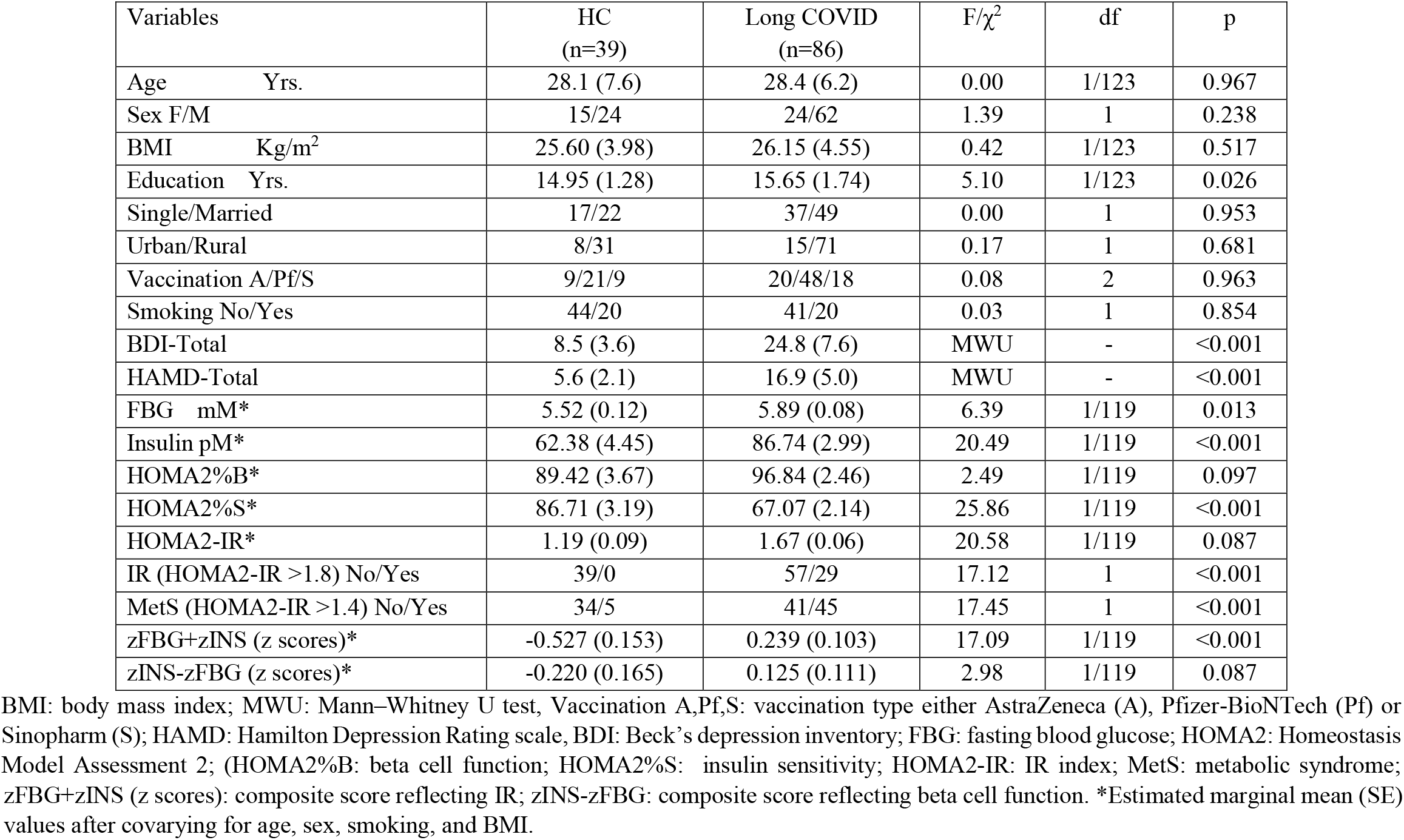
Sociodemographic characteristics and insulin resistance (IR) data in Long COVID patients and healthy controls (HC)

### Results of cluster analysis

In order to delineate a cluster of Long COVID patients characterized by disorders in biomarkers and increased depression scale scores, we performed K means factor analysis using OSTOX, zNLRP3+CRP, zFBG+zINS, and zBDI+zHAMD as clustering variables. Two clusters were formed with an adequate silhouette measure of cohesion and separation of 0.6, cluster 1 comprises 63 subjects, and cluster 2 comprises 61 subjects. Only Long COVID patients were allocated to cluster 2, while cluster 1 consists of 25 patients and 39 healthy controls. Table 2 demonstrates the sociodemographic data of Cluster 1 and Cluster 2 subjects. No significant differences between these study groups were detected in age, BMI, sex, vaccination type, and TUD, whereas subjects in cluster 2 have a significantly higher PBT, zPBT-zSpO2, and total BDI and HAMD scores, and lower SpO2-than Cluster 1 subjects. FBG and insulin, HOMA-2IR, and the zFBG+zINS score were significantly greater in cluster 2 than in cluster 1, whereas there were no significant differences in HOMA2%B, and zINS-zFBG. The HOMA2%S and zINS-zFBG indices were significantly lower in cluster 2 than in cluster 1. Covarying for age, BMI, sex, and TUD showed that none of these variables had any effect on the IR-related data. Table 2 shows that zOSTOX, AOPP, zNLRP3, zNLRP3+zCRP, CRP, caspase 1, IL-1β, and IL-18 were significantly higher in Cluster 2 as compared with cluster 1.

**Table 2.** Features of the two clusters generated using K mean cluster analysis

| **Variables** | **Cluster 1**<br>**N=64** | **Cluster 2**<br>**N=61** | **F/$\chi^2$** | **df** | **p** |
| --- | --- | --- | --- | --- | --- |
| Age Yrs. | 28.1 (7.2) | 28.59 (6.00) | 0.17 | 1/123 | 0.686 |
| Sex F/M | 22/42 | 17/44 | 0.62 | 1 | 0.433 |
| Healthy controls/patients | 39/25 | 0/61 | 54.03 | 1 | <0.001 |
| BMI Kg/m<sup>2</sup> | 25.66 (3.68) | 26.32 (5.01) | 0.70 | 1/123 | 0.404 |
| Vaccination A/Pf/S | 14/33/17 | 15/36/10 | 1.91 | 2 | 0.385 |
| Peak body temperature | 37.39 (0.75) | 38.75 (0.96) | 78.29 | 1/123 | <0.001 |
| Lowest SpO2 (%) | 93.58 (2.62) | 90.85 (4.56) | 16.98 | 1/123 | <0.001 |
| zPBT_zSPO2 | -0.532 (0.686) | 0.558 (0.977) | 52.57 | 1/123 | <0.001 |
| BDI-Total | 12.8 (7.5) | 26.87 (6.8) | 119.73 | 1/123 | <0.001 |
| HAMD-Total | 8.6 (4.7) | 18.33 (4.8) | 131.02 | 1/123 | <0.001 |
| FBG mM | 5.63 (0.09) | 5.92 (0.10) | 4.68 | 1/120 | 0.032 |
| Insulin pM | 70.91 (3.58) | 87.78 (3.67) | 10.77 | 1/120 | 0.001 |
| HOMA2%B | 92.33 (2.87) | 96.82 (2.94) | 1.19 | 1/120 | 0.278 |
| HOMA2%S | 80.28 (2.57) | 65.77 (2.63) | 15.43 | 1/120 | <0.001 |
| HOMA2 IR | 1.36 (0.07) | 1.69 (0.07) | 10.83 | 1/120 | 0.001 |
| zFBG+zInsulin (z scores) | -0.273 (0.122) | 0.286 (0.125) | 10.19 | 1/120 | 0.002 |
| zINS-zFBG (z scores) | 0.114 (0.133) | -0.075 (0.130) | 1.03 | 1/120 | 0.311 |
| AOPP (μmol/g) | 0.148 (0.113) | 1.1432 (0.116) | 4.30 | 1/120 | 0.040 |
| MPO (ng/ml) | 44.46 (2.57) | 51.28 (2.63) | 3.42 | 1/120 | 0.067 |
| zOSTOX (z scores) | -0.229 (0.123) | 0.267 (0.126) | 7.91 | 1/120 | 0.006 |
| CRP (mg/L) | 5.26 (0.43) | 8.36 (0.44) | 29.59 | 1/120 | <0.001 |
| Caspase1 (pg/ml) | 69.10 (2.56) | 81.98 (2.62) | 12.30 | 1/120 | <0.001 |
| IL-1β (pg/ml) | 4.40 (0.25) | 5.92 (0.25) | 18.37 | 1/120 | <0.001 |
| IL-18 (pg/ml) | 219.59 (9.22) | 257.75 (9.45) | 8.30 | 1/120 | 0.005 |
| zNLRP3 (z scores) | -0.496 (0.109) | 0.463 (0.112) | 37.40 | 1/120 | <0.001 |
| zNLRP3+zCRP (z scores) | -0.641 (0.096) | 0.673 (0.098) | 91.01 | 1/120 | <0.001 |
| zNeurotoxicity (z scores) | -1.958 (0.266) | 1.999 (0.272) | 107.71 | 1/119 | <0.001 |

BMI: body mass index; Vaccination A,Pf,S: vaccination type either AstraZeneca (A), Pfizer-BioNTech (Pf) or Sinopharm (S); SpO<sub>2</sub>: oxygen saturation; HAMD: Hamilton Depression Rating Scale; BDI: Beck Depression Inventory; FBG: fasting blood glucose; HOMA2: Homeostasis Model Assessment 2; HOMA2%B: beta cell function; HOMA2%S: insulin sensitivity; HOMA2-IR: insulin resistance index; MetS: metabolic syndrome; zFBG+zINS: composite score reflecting IR; zINS-zFBG: composite score reflecting beta cell function; AOPP: advanced oxidation protein products; MPO: myeloperoxidase; CRP: C-reactive protein, IL: interleukin, zOSTOX: composite score of oxidative stress, NLRP3: the NLRP3 inflammasome computed as zIL-1 $\beta$ +zIL-18+zcaspase1; zNeurotoxicity: a composite score based on zIR+zNLRP3+zCRP+zOSTOX.

### Intercorrelation matrix

**Table 3** shows the intercorrelation matrix between zFBG+zINS and zINS-zFBG and other clinical data and biomarkers. zFBG+zINS was significantly correlated with zBDI+zHAMD, total BDI, total HAMD, and PBT. There were no significant correlations between zFBG+zINS and zINS-zFBG, zNLRP3+zCRP, zOSTOX, or SpO2. There were no significant correlations between zINS-zFBG and any of the other variables.

**Table 3.**
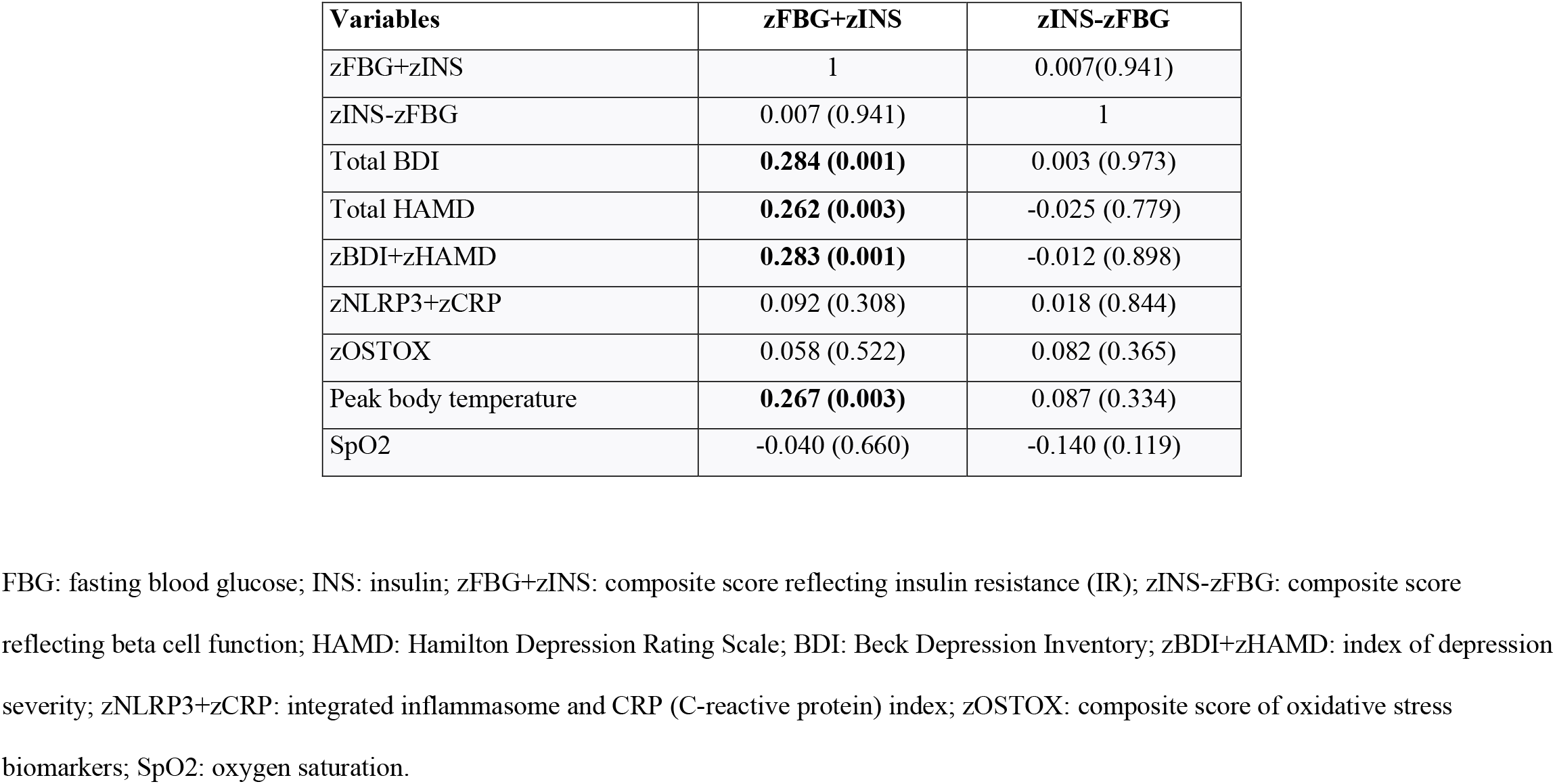
Intercorrelation matrix between insulin resistance (zFBG+zINS) and beta cell function (zINS-zFBG) and other biomarkers.

| **Variables** | **zFBG+zINS** | **zINS-zFBG** |
| --- | --- | --- |
| zFBG+zINS | 1 | 0.007(0.941) |
| zINS-zFBG | 0.007 (0.941) | 1 |
| Total BDI | **0.284 (0.001)** | 0.003 (0.973) |
| Total HAMD | **0.262 (0.003)** | -0.025 (0.779) |
| zBDI+zHAMD | **0.283 (0.001)** | -0.012 (0.898) |
| zNLRP3+zCRP | 0.092 (0.308) | 0.018 (0.844) |
| zOSTOX | 0.058 (0.522) | 0.082 (0.365) |
| Peak body temperature | **0.267 (0.003)** | 0.087 (0.334) |
| SpO2 | -0.040 (0.660) | -0.140 (0.119) |

FBG: fasting blood glucose; INS: insulin; zFBG+zINS: composite score reflecting insulin resistance (IR); zINS-zFBG: composite score reflecting beta cell function; HAMD: Hamilton Depression Rating Scale; BDI: Beck Depression Inventory; zBDI+zHAMD: index of depression severity; zNLRP3+zCRP: integrated inflammasome and CRP (C-reactive protein) index; zOSTOX: composite score of oxidative stress biomarkers; SpO2: oxygen saturation.

### Prediction of the scores of depression scales using biomarkers

**Table 4** shows the results of the multiple regression analysis with the total scores of the HAMD and BDI as dependent variables and the measured biomarker composite scores as explanatory variables while allowing for the effects of age, sex, education, TUD, and BMI. Regression #1 shows that zNLRP3+CRP, zFBG+zINS, and zOSTOX (all three significantly and positively associated) explained 26.4% of the variance in the zBDI+zHAMD score. **Figure 1** shows the partial regression plot of zBDI+zHAMD on zFBG+zINS after adjusting for the variables listed in Table 4. In regression #2, 56.8% of the variance in the zBDI+zHAMD score could be explained by PBT and zNeurotoxicity. **Figure 2** shows the partial regression plot of zBDI+zHAMD on zNeurotoxicity. We found that 24.8% of the variance in the total BDI score could be explained by the regression on zNLRP3+CRP, zFBG+zINS, and zOSTOX (Regression #3). Regression #4 shows that 24.7% of the variance in the total HAMD score could be explained by the regression on the zNLRP3+CRP, zFBG+zInsulin, and zOSTOX.

**Table 4.** Results of multiple regression analysis with depression scale scores as dependent variables and biomarkers as explanatory variables.

| Dependent Variables | Explanatory Variables | $\beta$ | t | p | F model | df | p | R<sup>2</sup> |
| --- | --- | --- | --- | --- | --- | --- | --- | --- |
| #1 zBDI+zHAMD | **Model** |  |  |  | 14.48 | 3/121 | <0.001 | 0.264 |
|  | zNLRP3+zCRP | 0.348 | 4.42 | <0.001 |  |  |  |  |
|  | zFBG+zINS | 0.238 | 3.04 | 0.003 |  |  |  |  |
|  | zOSTOX | 0.218 | 2.78 | 0.006 |  |  |  |  |
| #2. zBDI+zHAMD | **Model** |  |  |  | 80.26 | 2/122 | <0.001 | 0.568 |
|  | Peak body temperature | 0.632 | 9.33 | <0.001 |  |  |  |  |
|  | zNeurotoxicity | 0.207 | 3.06 | 0.003 |  |  |  |  |
| #3. Total BDI | **Model** |  |  |  | 13.287 | 3/121 | <0.001 | 0.248 |
|  | zNLRP3+zCRP | 0.307 | 3.85 | <0.001 |  |  |  |  |
|  | zFBG+zInsulin | 0.242 | 3.05 | 0.003 |  |  |  |  |
|  | zOSTOX | 0.242 | 3.04 | 0.003 |  |  |  |  |
| 4. Total HAMD | **Model** |  |  |  | 13.21 | 3/151 | <0.001 | 0.247 |
|  | zNLRP3+zCRP | 0.365 | 4.58 | <0.001 |  |  |  |  |
|  | zFBG+zINS | 0.218 | 2.75 | 0.007 |  |  |  |  |
|  | zOSTOX | 0.179 | 2.25 | 0.026 |  |  |  |  |

HAMD: Hamilton Depression Rating Scale, BDI: Beck Depression Inventory, zBDI+zHAMD: composite score comprising BDI and HAMD, zNLRP3+zCRP: composite score comprising the NLRP3 inflammasome and CRP (C-reactive protein), zOSTOX: composite of oxidative stress biomarkers, SpO2: peripheral oxygen saturation, FBG: fasting blood glucose, INS: insulin, zFBG-zINS: index of insulin resistance.

**Figure 1.**
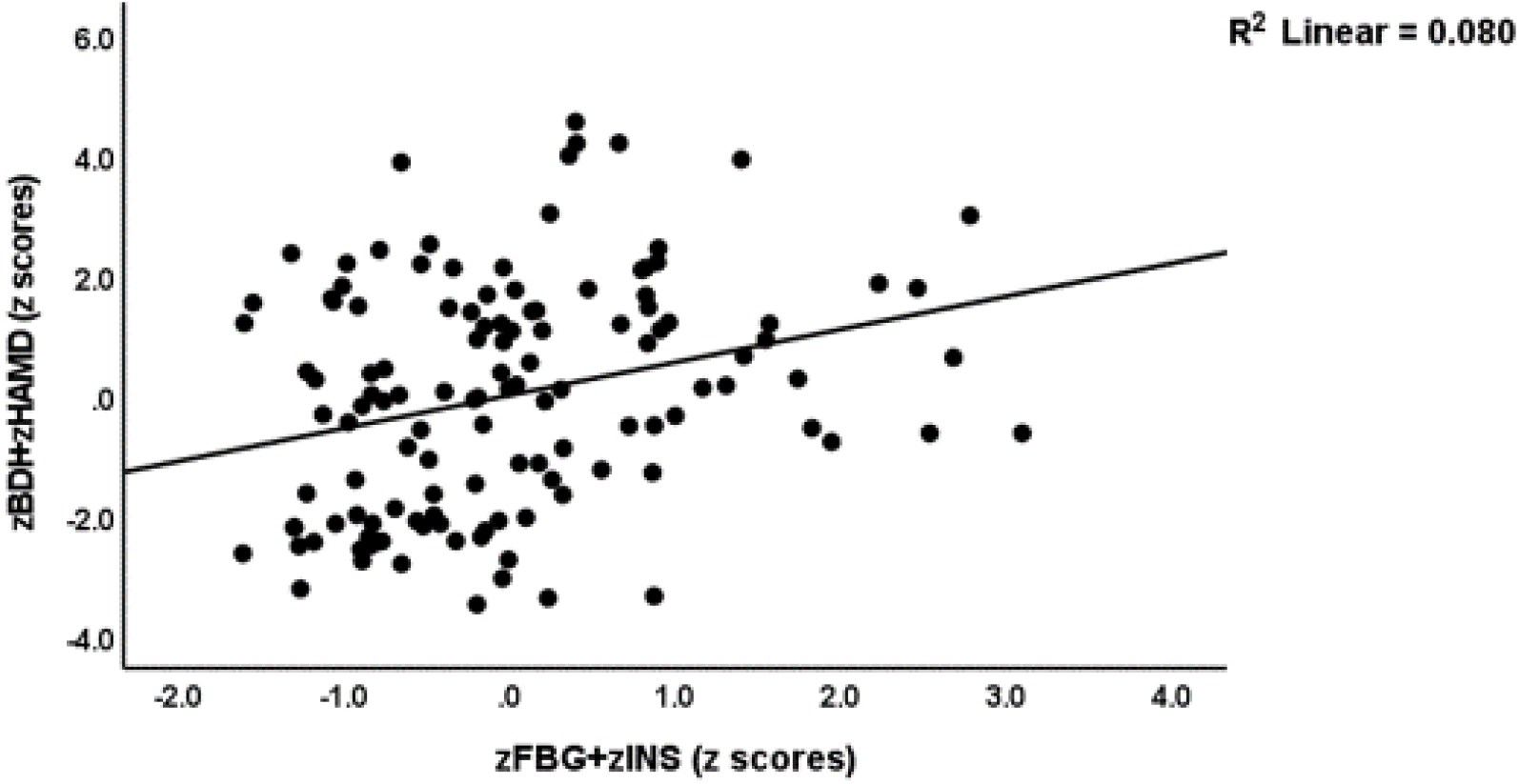
Partial regression plot of severity of depression (zBDl+zHAMD) on the index of insulin resistance (zFBG+zINS) BDI: Beck Depression Inventory: HAMD: Hamilton Depression Rating scale; FBG: fasting blood glucose; INS: insulin

**Figure 2.**
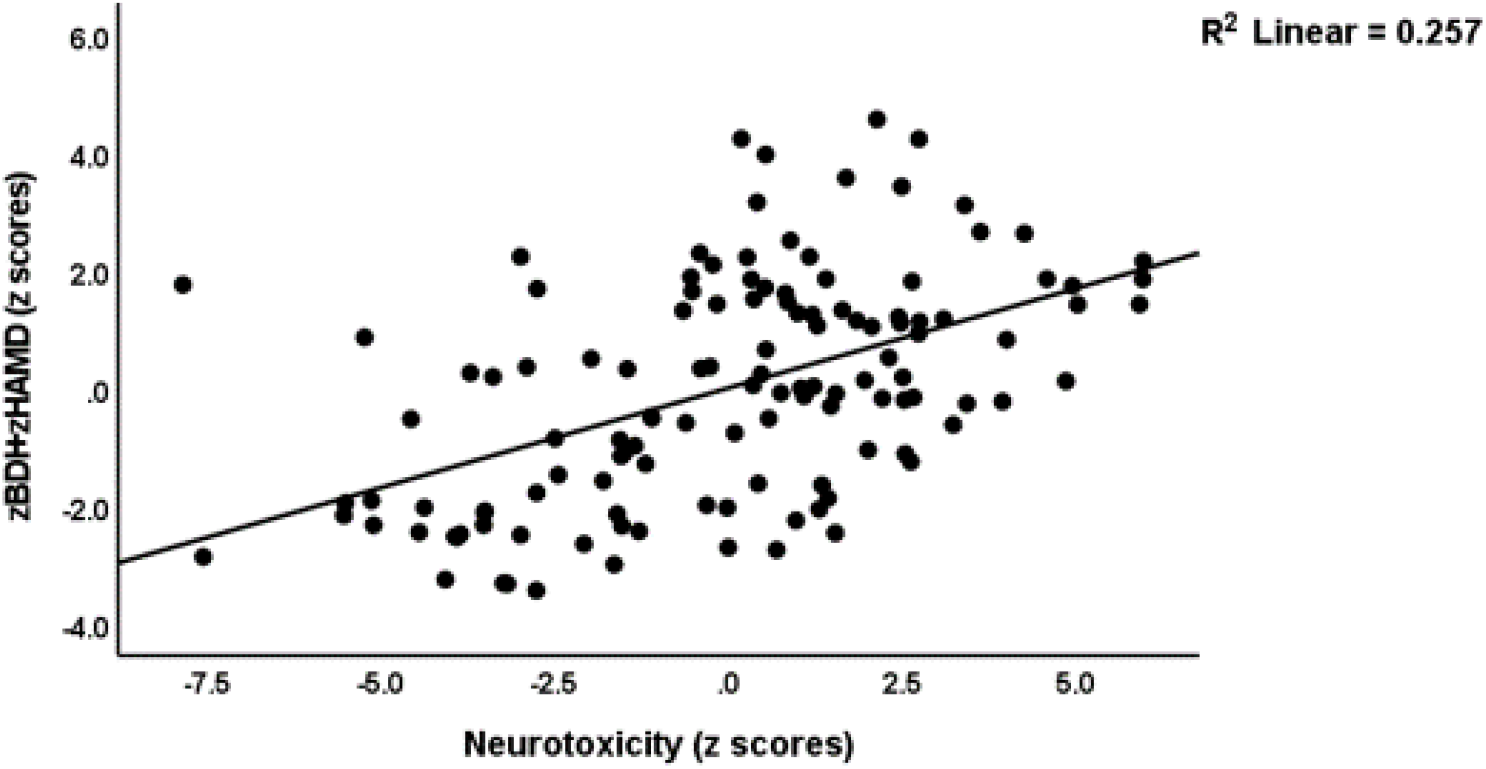
Partial regression plot of severity of depression (zBDT+zHAMD) on the index of neurotoxicity. BD1: Beck Depression Inventory; HAMD: Hamilton Depression Rating scale

### Results of PLS path and PLS predict analysis

**Figure 3** shows the final PLS model obtained after feature selection, prediction-oriented segmentation with multi-group analysis, and PLS prediction analysis. The model shows adequate quality fit data including an SRMR of 0.013. We found that 26.4% of the variance in depression severity (a factor extracted from the total HDRS and BDI scores) was explained by zFBG+zINS, OSTOX, and zNLRP+CRP. Increased PBT explained part of the variance in zFBG+zINS (7.1%), OSTOX (3.5%), and zNLRP+CRP (17.1%). PBT had a significant indirect effect on depression severity (t=4.49, p<0.001), which was mediated by zFBG+zINS (t=2.13, p=0.034) and zNLRP3+zCRP (t=2.86, p=0.004).

**Figure 3.**
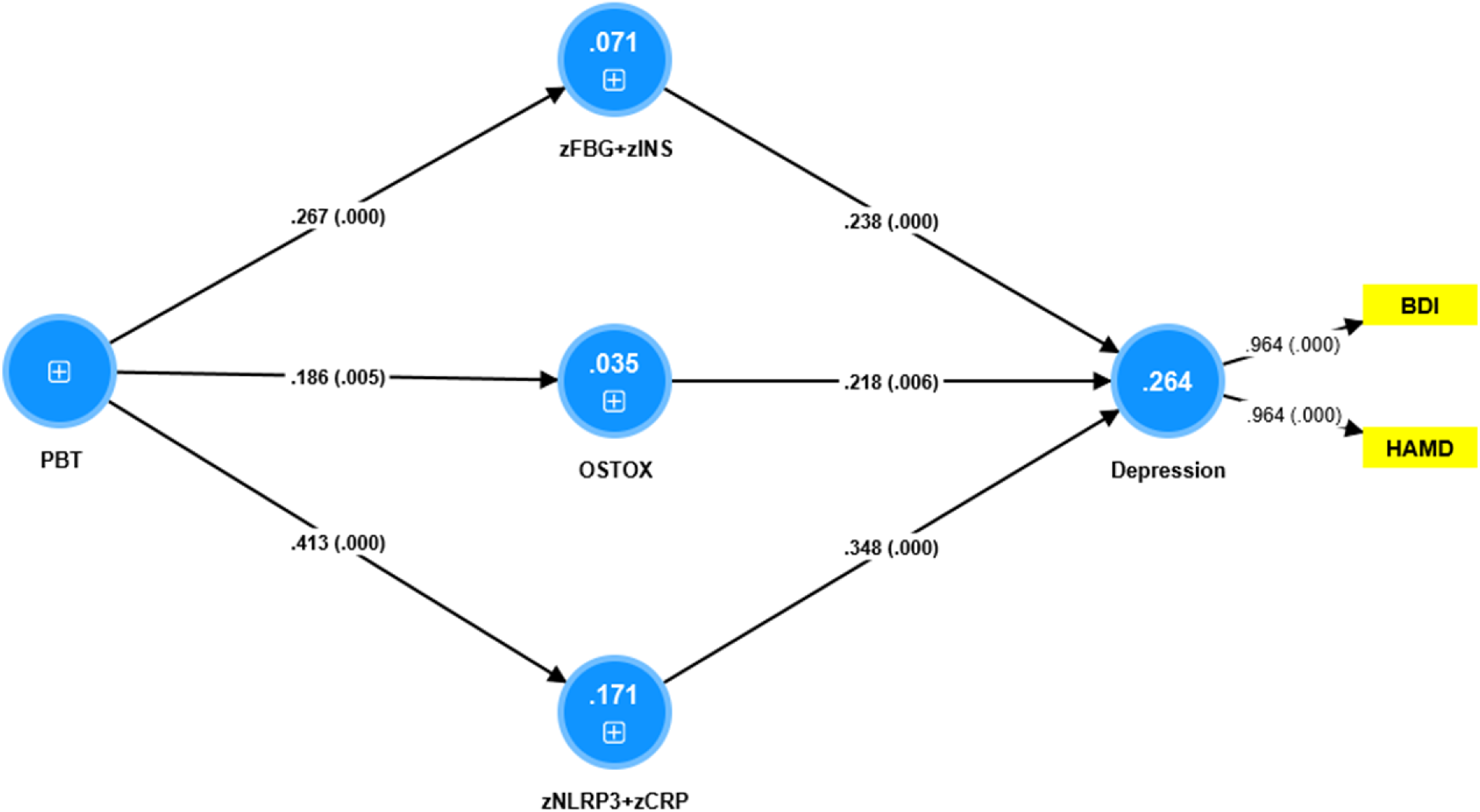
Results of partial regression analysis with severity of depression (entered as a latent vector) as dependent variable. Direct predictors are the index of insulin resistance (zFBG+zINS), index of oxidative stress toxicity (OSTOX) and an index of inflammation (zNLRP3+zCRP). Peak body temperature (PBT) during the acute phase of illness predicts the biomarkers of Long COVID. BDI: Beck Depression Inventory; HAMD: Hamilton Depression Rating scale; FBG: fasting blood glucose; INS: insulin; NLRP3: index of the nucleotide-binding domain, leucine-rich repeat, and pyrin domain-containing protein 3 inflammasome; CRP: C-reactive protein.

## Discussion

### Differences in IR parameters between Long-COVID and controls

The first major finding of the present study is that Long COVID is characterized by increased HOMA2-IR, FBG, insulin, and lowered HOMA2%S. Moreover, using threshold values of HOMA2-IR to denote IR (HOMA2-IR>1.8) and MetS (HOMA2-IR>1.4) ^(38)^, we found that 33.7% of the patients fulfilled the criteria for IR (versus 0% of the controls) and 52.3% for MetS (versus 12.8% of the normal controls). It is known that T1DM and T2DM patients are more vulnerable to SARS-CoV-2 and increased risk of critical disease and mortality due to COVID-19 ^(1, 41)^. Moreover, COVID-19 may increase glucose levels, aggravate IR, and cause new-onset IR and chronic metabolic disorders that did not exist before the COVID-19 infection ^(41-43)^. Our results of an increased HOMA2-IR index and increased IR in Long COVID patients extend previous results that the prevalence rate of IR ranges between 8.2% and 15% in hospitalized Chinese children and adults infected with SARS-CoV-2 ^(44, 45)^, whilst higher prevalence rates (34.6%) were observed in COVID-19 patients with the more severe and critical disease ^(45)^.

### Biomarkers of increased IR in Long COVID

The second major finding of this study is that IR due to Long COVID is predicted by increased PBT during the acute phase of infection. Contrary to the a priori hypothesis, IR was not significantly associated with lowered SpO2 during the acute phase and with key IO&NS biomarkers of Long COVID, such as increased CRP, NLRP3 activation, and oxidative pathways (including MPO and AOPP levels). The results of our PLS analysis show that the severity of the inflammatory response during the acute infectious phase is associated with increases in IR (small effect size of 7.1%), inflammatory responses (medium effect size of 17.1%), and oxidative toxicity (small effect size of 3.5%) during Long COVID.

The effects of the immune-inflammatory response during the acute phase on new-onset IR may be explained by the knowledge that SARS-CoV-2 may cause an (excessive) immune response with a release of a wide spectrum of cytokines generating a systemic proinflammatory milieu with increased levels of IL-6 and TNF-α which are known to contribute to IR, β-cell hyperstimulation, and, ultimately, β-cell death ^(41, 43, 46, 47, 48, 49)^. In addition, low-grade inflammation, as may be observed in Long COVID ^(17)^ is known to contribute to the development of IR and the progression to T2DM ^(50, 51)^, for example, through increases in gluconeogenic stress hormones and the resulting increased demand for β-cells ^(52)^.

Our negative findings that lowered SpO2 and increased MPO levels have no discernable effect on the HOMA2-IR index contrast with the knowledge that intermittent hypoxia can lead to increased β-cell proliferation and death due to oxidative stress ^(53)^. The pancreatic β-cell is metabolically very active and highly dependent on oxygen supply ^(54)^, whilst β-cells have relatively low antioxidant levels and are, therefore, very sensitive to oxidative stress ^(55-57)^. In addition, hypoxia, followed by reoxygenation, elicits oxidative stress ^(58)^. Our negative findings that increased O&NS, including MPO levels, have no significant impact on the HOMA2-IR index contrast with the knowledge that COVID-19-associated elevations in MPO are associated with the HOMA2-IR index ^(42)^. Moreover, treatment with MPO may upregulate gluconeogenesis, TNF-α, and IL-6 gene expression in adipocytes and HUVEC cells, resulting in increased IR ^(42)^. Although increased reactive oxygen species impact insulin receptor signaling, thereby, contributing to IR ^(59)^, we could not detect any effects of O&NS on IR in Long COVID.

All in all, there is only a small effect of inflammation during the acute phase on new-onset IR, whilst activated IO&NS pathways during Long COVID appear to have no effect at all. This indicates that other factors related to the SARS-CoV-2 virus or COVID-19 are largely involved in new-onset IR. In this respect, evidence suggests that SARS-CoV-2 can infect and replicate in insulin-producing pancreatic β-cells, thereby resulting in impaired production and secretion of insulin and the metabolic dysregulation observed in patients with COVID-19 ^(60-63)^. Indeed, transcriptome analysis of infected pancreatic cell cultures confirmed that SARS-CoV-2 hijacks the ribosomal machinery in these cells ^(64)^. Microvascular damage caused by SARS-CoV-2 or via micro-thrombotic lesions may result in perfusion anomalies in pancreatic islets, which are required for glucose sensing and insulin release, whilst abnormal capillary architecture and fragmentation contribute to β-cell failure in T1DM and T2DM ^(65, 66)^. As such, SARS-CoV-2 infection may cause direct pancreatic injury, which may worsen existing IR in T1DM/T2DM or contribute to new-onset IR and T2DM ^(67, 68)^. Mechanistic explanations associated with COVID-19 are: a) COVID-19 is accompanied by a hypercoagulable state ^(69, 70)^ which may cause endothelial injuries leading to microvascular inflammation and thrombosis ^(71)^; b) autonomic dysfunction or autoimmunity ^(60, 61)^; and c) secondary mitochondrial dysfunctions ^(72)^.

### Insulin resistance and depression in Long COVID

The third major finding of this study is that the increased depression scores during Long COVID are significantly related to IR and that the latter impacts depression above and beyond the effects of inflammation, NLRP3 and O&NS activation. Previously, we have reviewed that there is a link between major depression and increased IR, T1DM/T2DM, and obesity ^(26, 27, 30)^. Apart from IO&NS processes, other mechanisms may explain this relationship including changes in the endocannabinoid system, mitochondrial dysfunctions, the gut microbiome, and the endogenous opioid system ^(30)^.

Our results suggest that IR in Long COVID may contribute to the overall neurotoxicity which comprises effects of IO&NS mechanisms ^(25, 73)^. Recently, we have described the mechanism that may explain these IR-associated neurotoxic effects on the physio-affective phenomenon of MDD ^(74)^. For example, IR is associated with a decrease in metabolic activity in the medial prefrontal cortex and hippocampal volume, dysfunctions in the medial prefrontal cortex and hippocampal connectome, and neurocognitive impairments including in executive functions and memory ^(75-77)^. Increased IR in the central nervous system is associated with lower levels of brain-derived neurotrophic factor, impaired synaptic plasticity, fewer dendritic spines, and neurodegeneration ^(78, 79)^. Furthermore, the endothelial dysfunctions caused by IR may result in blood-brain barrier breakdown, which in turn may lead to elevated translocation of proinflammatory compounds from the plasma compartment to the CNS, thereby contributing to neuronal abnormalities, neuroinflammation, and neurodegeneration ^(80, 81)^.

### Limitations

The results of this study would have been more informative if we had measured atherogenicity biomarkers, gluconeogenic stress, and MetS-related peptide hormones including adiponectin, leptin, ghrelin and somatostatin, and mitochondrial dysfunctions.

## Conclusions

The results show that long COVID is associated with increased IR and new onset IR (based on a HOMA2-IR INDEX > 1.8) and that the increased IR is only mildly associated with signs of the inflammatory response during the acute phase and not at all with the mild IO&NS processes during Long COVID. IR due to Long COVID is associated with depressive symptoms that are present in Long COVID and increased IR may contribute to the neurotoxicity pathophysiology of depression together with IO&NS pathways. Future research should examine other possible mechanisms leading to IR in Long COVID including hypercoagulation, endothelial injuries, microvascular inflammation, thrombosis, and mitochondrial functions during acute COVID-19 infection, and the onset of autoimmune responses in Long COVID. One possibility could be to evaluate treatments with metformin which may be useful to treat depression ^(82)^ and improving the outcome of COVID-19 patients and patients with pre-existing T2DM ^(83, 84)^.

## Data Availability

To avoid plagiarism, the dataset generated during and/or analysed during the study are not publicly available but are available from the corresponding author on reasonable request once the dataset is fully exploited by the authors.

## Author’s contributions

All authors contributed substantially to the conceptualization of the analysis, the interpretation of the results, the writing and editing of the paper, and the final version of the manuscript.

## Ethics approval and consent to participate

Before participating in the study, all controls and patients, or their parents or legal guardians, supplied written informed consent. The Najaf Health Directorate, Training and the Human Development Center (Document No. 18378/2021) and the institutional ethics boards of the University of Kufa (8241/2021) both gave their approval for the study.

## Acknowledgments

We appreciate the assistance of the workers at Al-Sadr Teaching Hospital and Al-Amal Specialized Hospital for Communicable Diseases in the Najaf governorate of Iraq. For their assistance in determining biomarker levels, we’d also want to thank the highly-skilled employees of the hospital’s internal laboratories.

